# ViralSim - A Geographically Informed Viral Transmission Simulation with Phylogeny-Based Fitness Analysis

**DOI:** 10.1101/2025.07.07.25330516

**Authors:** Benjamin Cai

## Abstract

Accurately simulating viral evolution is critical for evaluating analytical tools, forecasting outbreaks, and improving our understanding of pathogen dynamics. ViralSim introduces a geographically structured agent-based model that emphasizes spatial transmission across clustered populations. Unlike prior approaches that primarily focus on genetic, purely statistical, or temporal parameters, ViralSim incorporates location-driven spread and explicit trait evolution. More over, the produced phylogeny enables the calculation and fine-tuning of diverse fitness metrics directly from ViralSim. These fitness measures provide insight into lineage success, and with enough development, offer a new way to analyze phylogenies. By combining spatial transmission, trait development, and fitness evaluation, ViralSim enables controlled benchmarking of phylogenetic and epidemiological analyses.

## 1 Introduction

Viral simulations constitute a critical methodological framework for modeling the transmission, mutation, and control of viral pathogens using computational and statistical techniques. These simulations enable the exploration of epidemic dynamics, inform the design of public health interventions, and support the forecasting of pandemic trajectories. Importantly, they allow researchers to investigate parameters and scenarios that are infeasible or unethical to assess empirically. Prior simulation efforts have concentrated on specific pathogens such as SARS-CoV-2, HIV, and influenza [1, 2, 3], incorporating complex biological processes such as gene-level mutation [4], diverse mutation models [5], and recombination events [6].

Given the constrained scope of this project, limited computational resources, and the early stage of technical expertise of the author, `ViralSim` focuses specifically on simulating the emergence and propagation of geographically localized transmission clusters. Although viral spread occurs at the level of individuals, larger-scale patterns emerge across social units—ranging from households and workplaces to cities and nations. While coalescent-based models have historically dominated viral phylogenetic simulations, an agent-based modeling (ABM) framework was selected due to its ability to more explicitly capture spatial heterogeneity and complex social dynamics. Though the comparative computational efficiency between coalescent and ABM approaches in forward-time simulations remains questionable in favor of ABMs, ABMs do offer distinct advantages in modeling nonlinear interactions and producing interpretable visualizations.

The primary objectives of `ViralSim` are threefold: (1) to explore conditions conducive to the emergence of pandemics, (2) to test the efficacy of various mitigation strategies, and (3) to evaluate and compare phylogenetic analysis methodologies across simulated datasets.

## 2 Approach

In `ViralSim`, the simulation framework is structured around three primary entities: *locations, hosts*, and *viruses*. These components interact over time to simulate the dynamics of viral transmission and evolution in a spatially structured population. Their roles are detailed as follows:

### 1. Locations

Each location represents a geographic or social unit. Locations may encode behavioral modifiers and structural interventions. For instance:

- A location might have a quarantine flag enabled, reducing the probability of transmission by a fixed percentage.
- A location can include periodic events (e.g., mass gatherings every 100 time steps), influencing host interactions and infection probability.
- All such parameters are configurable via input files, and for more specific behaviors, configurable based on instructions from the README in the codebase.

### 2. Hosts

Hosts represent individuals in the simulated population. With respect to any given viral family, each host exists in one of four states:

- *Susceptible*
- *Infected*
- *Asymptomatic*
- *Removed* (via death or acquired immunity)
- Removed hosts no longer participate in transmission for that specific viral family.

### 3. Viruses

Each virus object includes a unique sequence of nucleotide base pairs and a set of phenotypic traits influencing its transmissibility and behavior. Notably:

- infectionRate
- asymptomaticInfectionRate
- deathRate
- recoveryRate
- mutationRate

Additionally, each viral family maintains a dynamic phylogeny representing its evolutionary history. Inside of `ViralSim`, this phylogeny is represented by a newick string instead of a tree object due to memory constraints.

### Transmission Model

Unlike traditional approaches which define transmission through a fixed probability distribution, `ViralSim` uses an agent-based method. Transmission occurs when an infected or asymptomatic host encounters a susceptible host, with success determined by the infecting virus’s specific traits. Each infection carries a probabilistic chance of inducing mutation during replication.

### Mutation Dynamics

The model incorporates the following types of mutations:

- *Substitutions*
- *Insertions and deletions (indels)*
- *Recombination*

The relative frequencies of these events are user-definable. Typically, the following hierarchy holds:

substitutionRate *≫* indelRate

*≫* recombinationRate

Each mutation event results in a random change in the virus’s traits. These traits are interconnected through a balancing equation of the form:

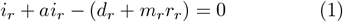

where:

- *i*_*r*_ = infection rate
- *ai*_*r*_ = asymptomatic infection rate
- *d*_*r*_ = death rate
- *r*_*r*_ = recovery rate
- *m*_*r*_ = mutation rate

At each mutation event, one trait is randomly selected to satisfy this constraint, while the others are shifted stochastically. Equation 1 is modifiable in ViralSim/src/model/Virus.cpp.

### Interaction and Mobility

Host movement occurs either according to predefined schedules or randomly (dependent on simulation scale and computational resources). The number of hosts to transition between locations is given as rate *m* times the total number of hosts, enabling spatial mixing. Over time, viruses accumulate mutations within hosts; if mutations exceeds a threshold *X*, immunity is bypassed and a new viral family is instantiated. Additionally, bypassing immunity events also triggers larger stochastic changes across these trait parameters, producing biologically unrealistic values, such as negative death rates which mirrors how some real-world viruses have extremely low if not a 0% death rate. This mechanism allows for lineage diversification and recurrent infection cycles.

## 3 Inputs and Outputs for ViralSim

The primary input mechanism for `ViralSim` is a structured command file named `commands.txt`. This file begins with a single integer specifying the number of simulation iterations, followed by a list of commands. Each command is of the format:

TimeStep CommandName [Arguments]

where `TimeStep` indicates when the command should be executed. The simulation parameters mentioned prior such as *iterations* (number of timesteps to simulate over), *m* (host location transition rate), and *X* (mutations to bypass immunity) are provided in a line before the first command in the form:

`iterations m X`

These inputs drive the stochastic agent-based dynamics of the simulation.

Below is a summary of supported and possibly supported commands:

| Command Name | Arguments | Effect |
| --- | --- | --- |
| AddVirus | Location, Host, InfectionRate ($\beta$), DeathRate ($\delta$), RecoveryRate ($\gamma$), MutationRate ($m$) | Introduces a virus with specified parameters to a host at the given location |
| AddLocation | Length, Width, Height, Number of Hosts ($X$), Max Hosts | Initializes a spatial location with dimensional constraints and host parameters |
| Masking (not implemented yet though configurable in code) | Location, Decrease | Reduces $\beta$ by Decrease for all hosts at specified location |
| Quarantine (not implemented yet though configurable in code) | Location, Time | Isolates the specified location, prohibiting host movement for the duration of Time |
| if (not implemented yet) | Condition, Command, Location | Executes Command if Condition (e.g., noVirus, lowHost) is satisfied at the given location |

`ViralSim` produces three output files for each simulated viral family: a multifasta sequence file, a corresponding metadata csv file, and a true phylogeny newick file. The sequence file contains the complete set of nucleotide sequences for all individual mutations within the viral lineage. The metadata file provides detailed annotations for each sequence, including its associated traits and the simulation time step at which it was sampled (note: stochastic or time-distributed sampling is currently only available through manual configuration in the codebase). The phylogeny file captures the full evolutionary history of the viral family, representing the ground truth against which phylogenetic inference methods may be benchmarked.

A notable feature in Figure 4 is the presence of clearly delineated clades. Three distinct groupings emerge, resembling the clade structures observed in real-world viral evolution, like the emergence of genetically differentiated SARS-CoV-2 ‘strains’.

**Figure 1:**
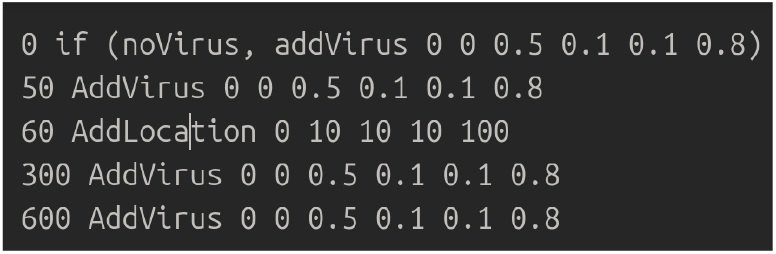
Example input commands.txt (lacking the simulation parameters)

**Figure 2:**
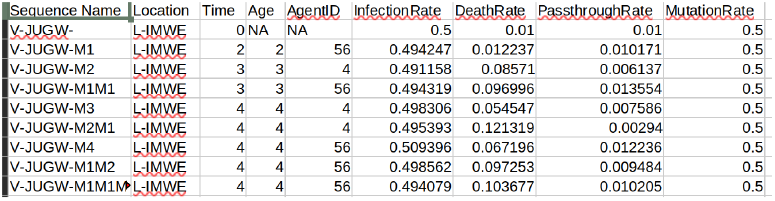
Example output metadata.

**Figure 3:**
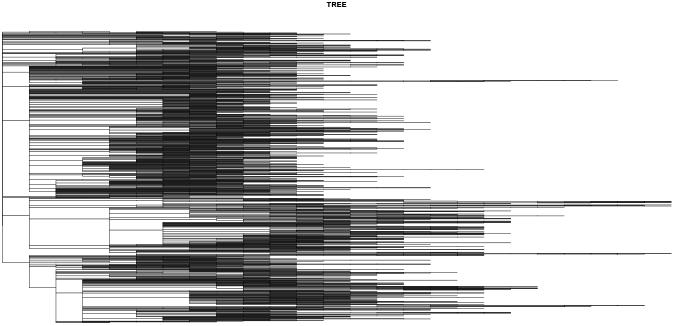
Example output phylogeny.

**Figure 4:**
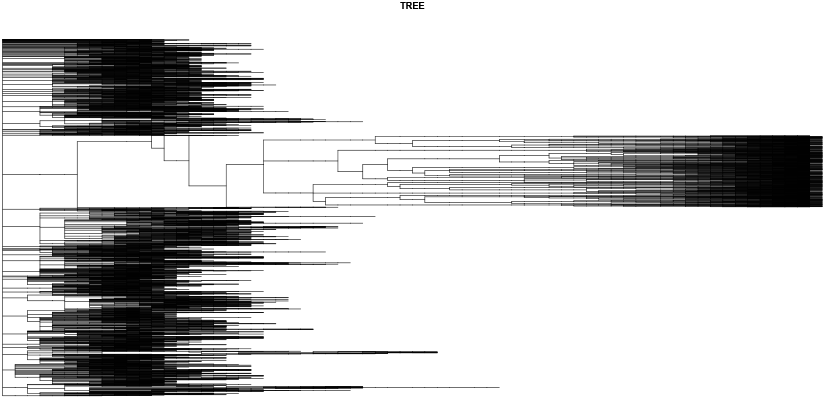
Another example output phylogeny.

## 4 Annotating Phylogenies by Geographic Origin

To extract important insights from simulated viral evolution, it is valuable to annotate phylogenetic trees with geographic information. One effective strategy is to identify and label transmission clusters—groups of genetically similar viral sequences that co-occur within the same geographic area (e.g., communities, cities, or countries).

Transmission clusters can be inferred by analyzing the distribution of sampled sequences at internal nodes of the phylogeny. Specifically, a node can be designated as the root of a transmission cluster if the majority of its descendants originate from a single location. The annotation procedure follows these steps:

1. For each internal node *P* (i.e., a node with at least one child), collect all descendant leaf nodes {*C*_1_, …, *C*_*k*_}.
2. For a given location *A*, compute the ratio of descendants belonging to location *A* versus those not from *A*.
3. If this ratio exceeds a predefined threshold (e.g., 100:1), label node *P* as the root of a transmission cluster associated with location *A*.
4. Repeat the process for all internal nodes and all candidate locations.

This procedure can be formalized using the following expression:

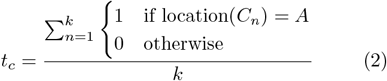

where:

- *t*_*c*_ is the proportion of child nodes assigned to location *A*,
- *k* is the total number of child nodes under the parent node *P*,
- location(*C*_*n*_) is the geographic origin of the *n*^th^ child.

Two metrics are critical when determining whether to annotate a node as a transmission cluster:

- *t*_*cr*_: the ratio of location-specific sequences (*t*_*c*_), which reflects location purity,
- *t*_*cn*_: the raw count of descendant sequences, indicating cluster size.

Nodes with low *t*_*cr*_ suggest spatial heterogeneity and should not be considered clusters. Conversely, nodes with low *t*_*cn*_ lack statistical significance. Both thresholds should be tuned depending on the sampling density and intended resolution.

## 5 Example Annotations on Real-World Data

Figure 5 demonstrates the application of the transmission cluster identification algorithm on real-world SARS-CoV-2 genomic data. Nodes within the phylogeny were marked if *t*_*cn*_ *>* 5 while opacity is based on *t*_*cr*_, revealing time and clade based transmission clusters.

**Figure 5:**
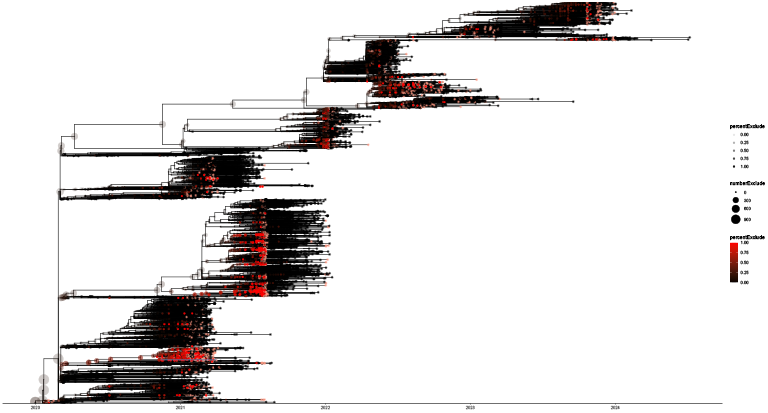
US SARS-CoV-2 Phylogeny annotated for California transmission clusters.

Temporal annotation, as shown in Figure 6, provides further insight into the chronological unfolding of transmission events. With a cluster ratio threshold of 30% and a minimum size of 5 descendants, the histogram above captures how geographically dominant clades emerge and fade over time. The peaks in cluster emergence can be found to correspond to public health choices and key events in the SARS-CoV-2 pandemic. This method facilitates retrospective understanding of regional spread dynamics, and, while not analyzed thoroughly in this study, might hold real-time and real-world capabilities/uses.

**Figure 6:**
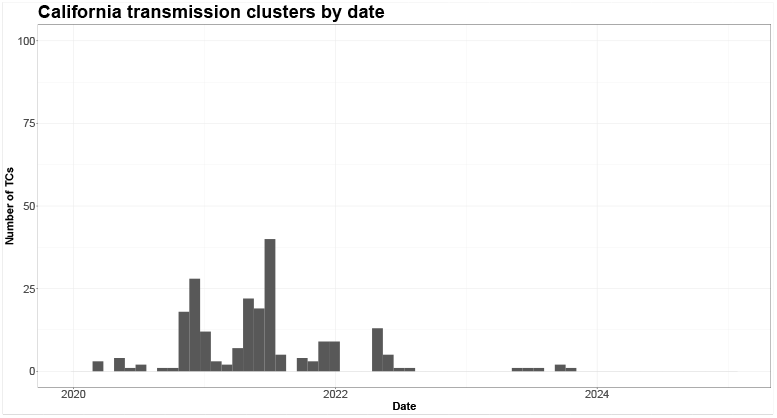
SARS-CoV-2 California Transmission Clusters through time with a t_cr_ = 30% and t_cn_ = 5.

## 6 Example Annotations on Simulated Data

Figures 7 and 8 display example annotations on simulated datasets generated by ViralSim. In both examples, clusters are visibly distinct, with subtrees dominated by sequences from a single location. These annotations serve as a check for simulation realism. More succinctly, the presence of clear, well-separated transmission clusters in the simulated data affirms that the simulation model successfully captures real-world viral spread. More over, the consistency between simulated and real-world clustering patterns supports the utility of this algorithm for transmission cluster phylogenetic annotation.

**Figure 7:**
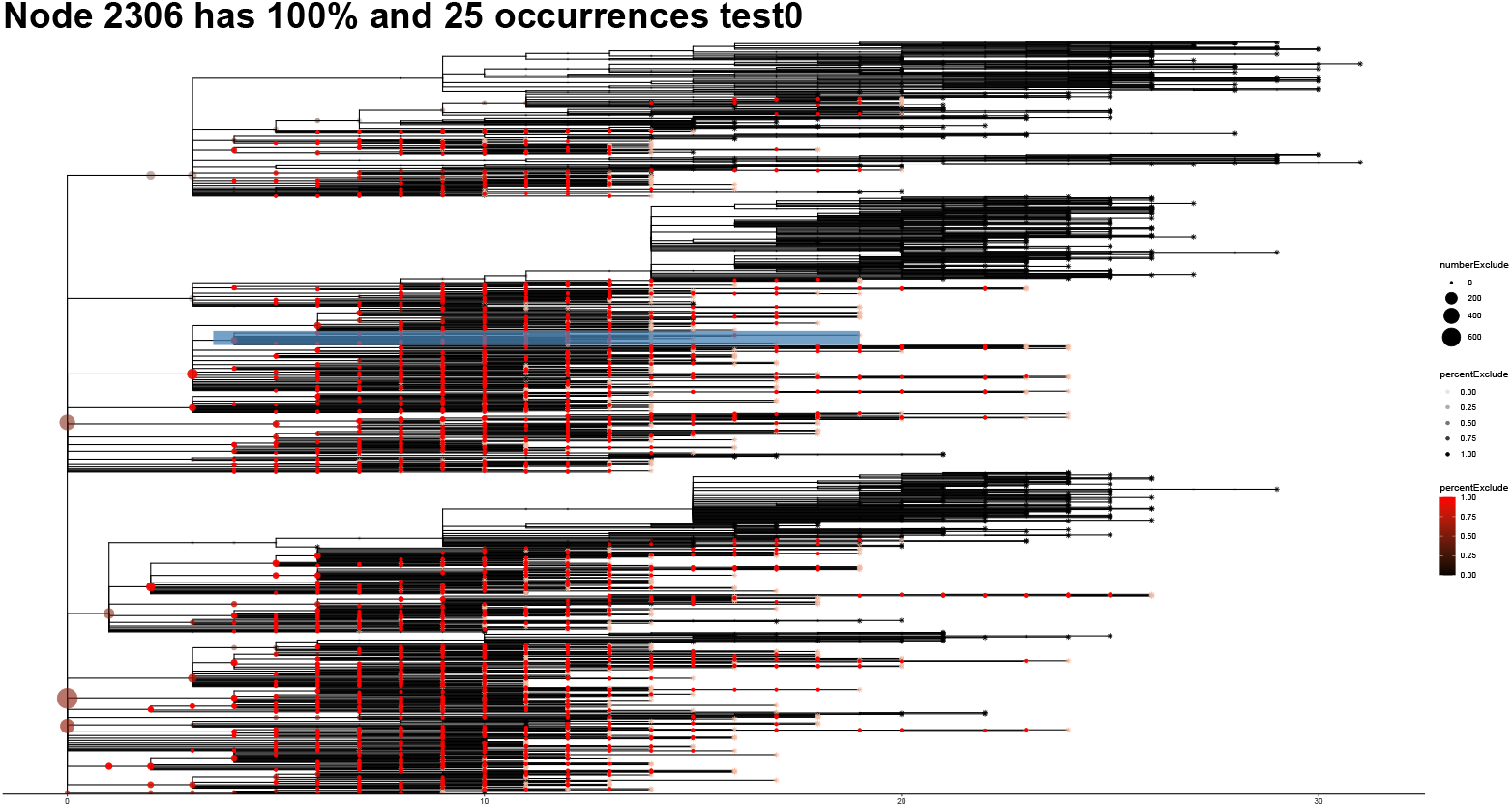
One example annotation on simulated data for location test0.

**Figure 8:**
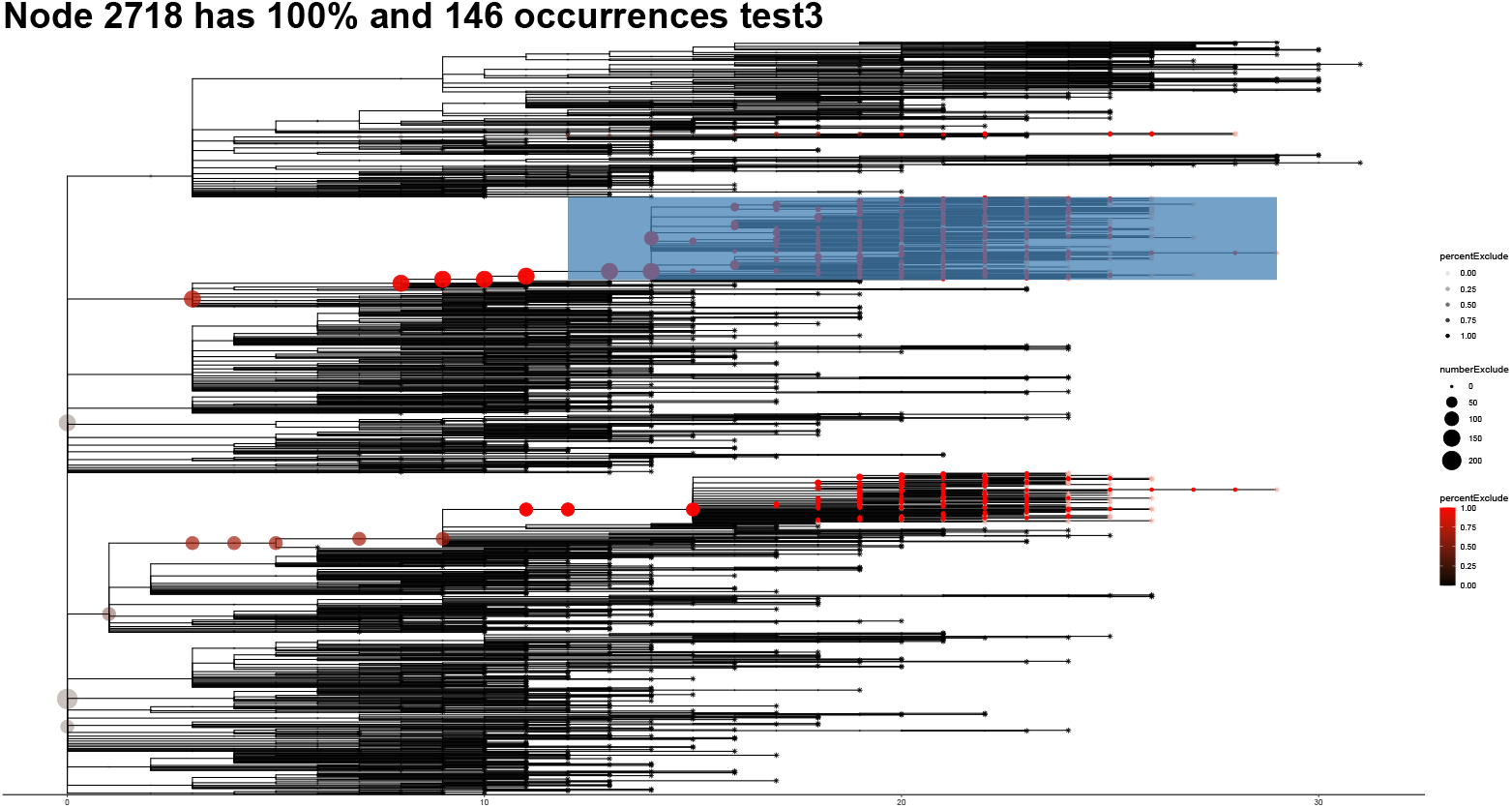
One example annotation on simulated data for location test3.

## 7 Evaluation Using Empirical Reconstruction Methods

Simulated sequence data enable benchmarking of phylogenetic reconstruction techniques. For instance, ultrafast methods like UShER [7] achieve high throughput on large datasets but may sacrifice some accuracy.

Unfortunately, currently, only maximum likelihood reconstruction was assessed.

Visual comparison between Figures 9 and 7, as well as Figures 10 and 8, indicates close correspondence in overall topology. Critically, major transmission cluster annotations align consistently. Minor rearrangements of clades, such as branch swapping, do not significantly alter topology or cluster interpretation.

**Figure 9:**
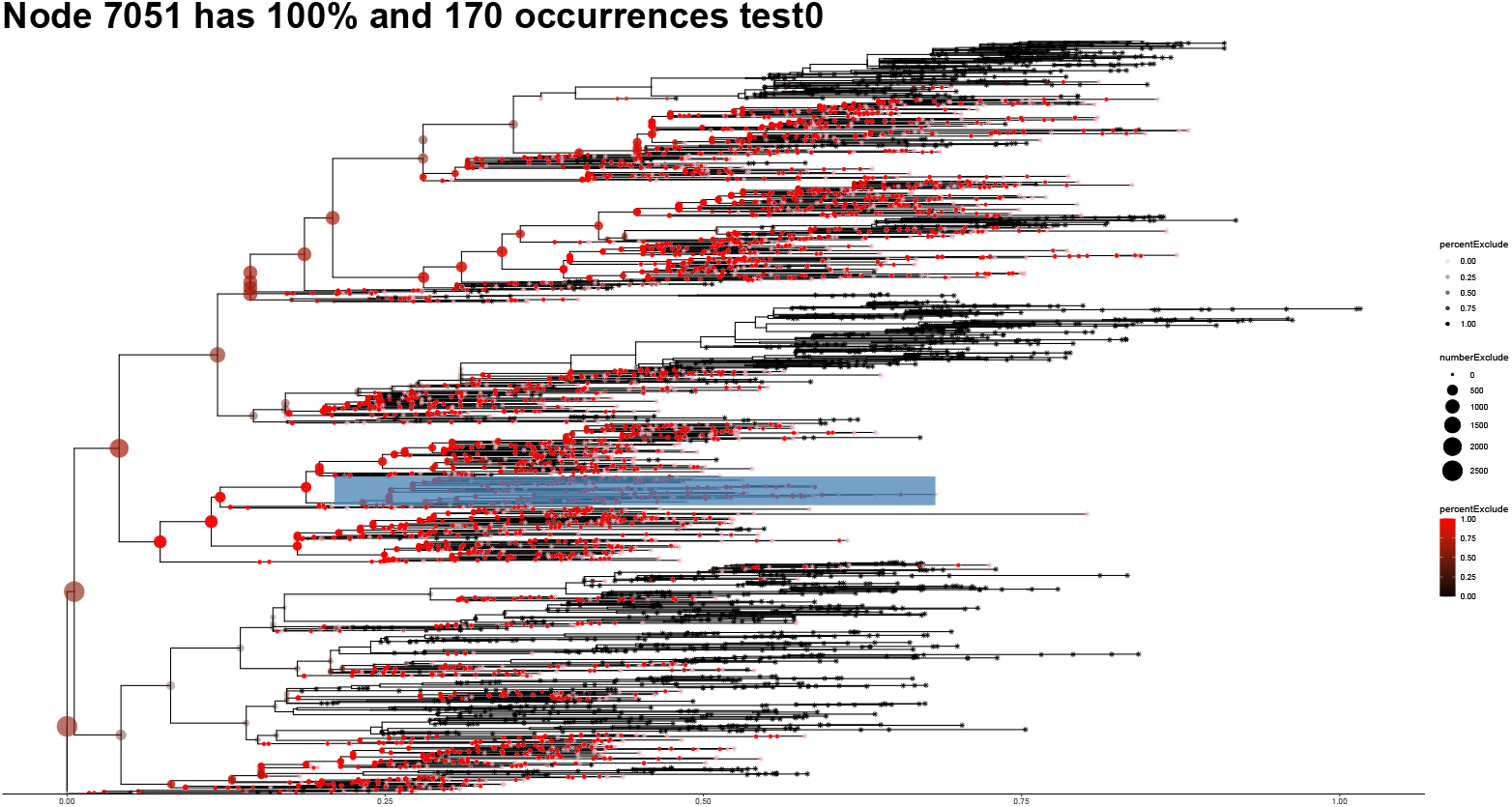
Annotation of a maximum likelihood phylogeny reconstructed from simulated data at location test0.

**Figure 10:**
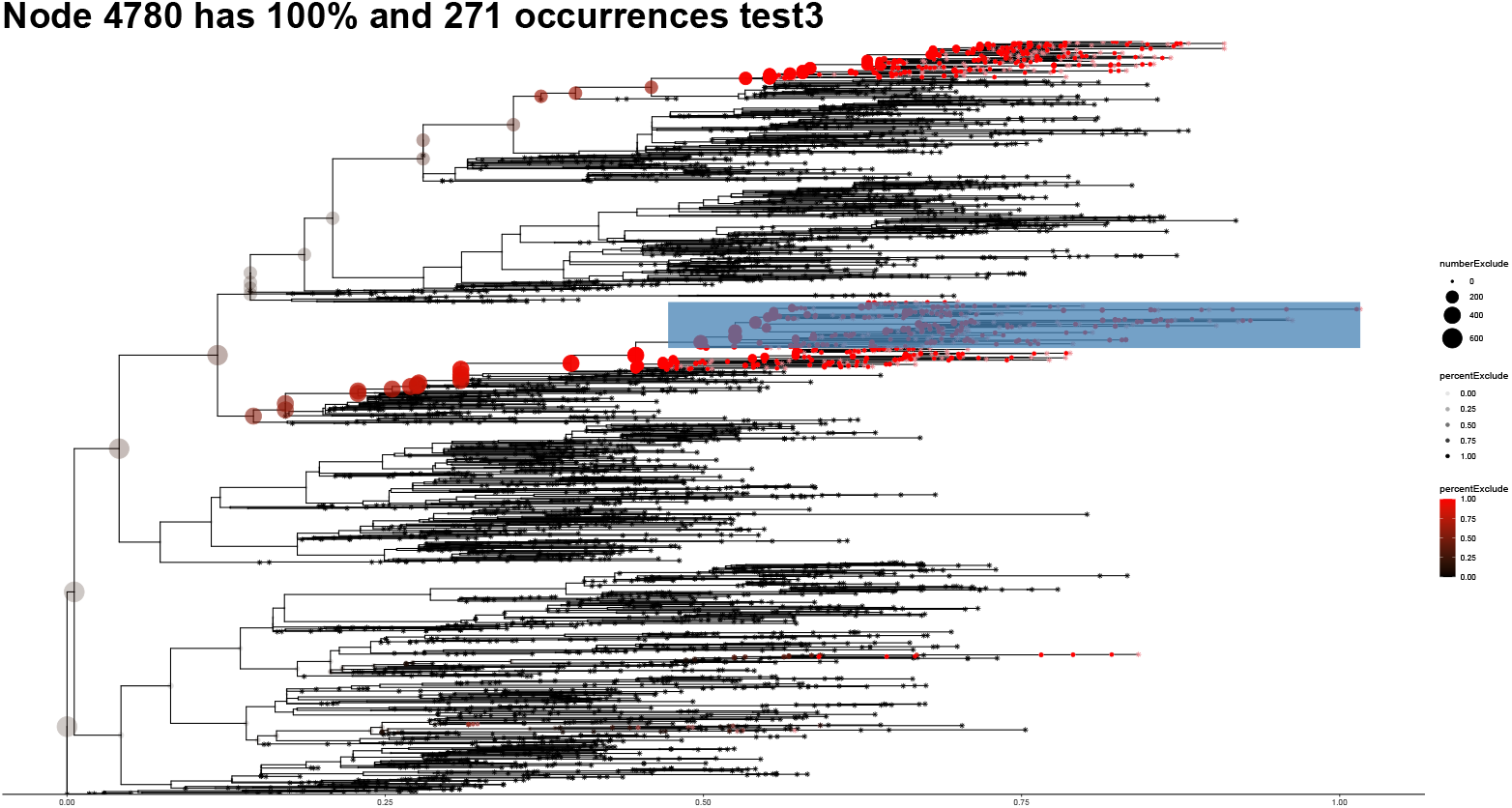
Annotation of a maximum likelihood phylogeny reconstructed from simulated data at location test3.

**Figure 11:**
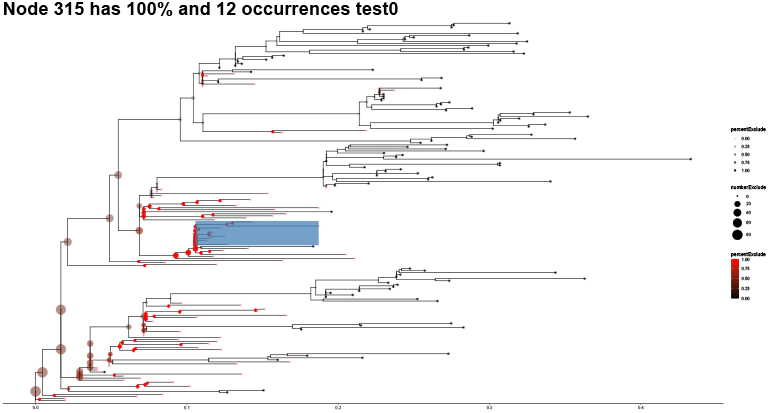
Annotation on a maximum likelihood phylogeny reconstructed from 20% sampled simulated data at location test0.

**Figure 12:**
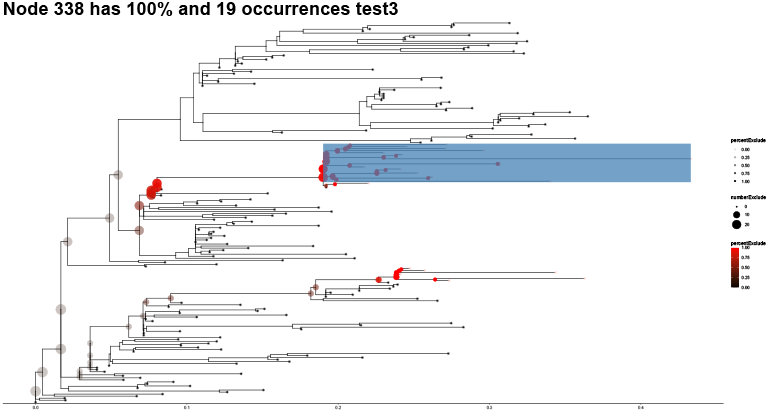
Annotation on a maximum likelihood phylogeny reconstructed from 20% sampled simulated data at location test3.

The discretization observed in time steps within simulated phylogenies results from the balance of computational load and event resolution per step, which does suggest potential for refinement by adjusting the granularity of actions per time step.

Furthermore, the impact of sampling bias on tree topology was investigated. Real-world viral phylogenetics rarely capture all mutations due to intrahost viral diversity and incomplete case reporting or sampling. Assessing phylogenetic robustness under partial sampling is therefore essential.

Despite reduced sampling, phylogenetic topology and transmission cluster integrity remain largely preserved. Variations such as branch rearrangements do not materially affect cluster inference, confirming the resilience of reconstructed trees to sampling limitations.

## 8 Fitness

Fitness (*F*) quantifies the spreading success of a virus node within a phylogenetic tree. One way that fitness can be defined is:

- **Terminal branch fitness:**

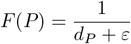

where *d*_*P*_ is the distance of node *P* to its parent.

- *Benefits:* Computationally simple for large number of nodes.
- *Drawbacks:* Ignores descendants and extremely sensitive to noise in reconstructed phylogenies (branch length estimation).

- **Subtree size fitness:**

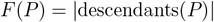

representing the count of all descendants of node *P* .

- *Benefits:* Captures reproductive success.
- *Drawbacks:* Does not reflect speed of diversification, also can place undue emphasis on root nodes.

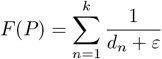

where:

- *F* (*P*) is the fitness of the node *P* .
- *k* is the number of descendents for node *P* including non-direct children.
- *d*_*n*_ is the branch length (distance) from the node *P* to its *n*^th^ descendent.
- *ε* is a tiny constant (1*e*6) included to avoid division by zero

### Interpretation

- A greater number of children (*k*) reflects higher fitness.
- Each child’s contribution decreases inversely with 10 branch length *d* .
- Shorter branch lengths imply stronger influence of parental traits on the child’s success.

### Computation for entire phylogeny

1. For node *P* (start at root and traverse tree), identify its children {*C*_1_, *C*_2_, …, *C*_*k*_}.
2. Measure branch lengths *d*_*n*_ from *P* to each child *C*_*n*_.
3. Compute each child’s contribution as 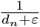.
4. Sum all contributions to obtain *F* .
5. Assign fitness *F* to node *P* .
6. Repeat for all nodes in the tree.

Within analysis of `ViralSim` outputs, several fitness metrics were separately implemented. Each one notes unique drawbacks and benefits which are listed here:

- **Local imbalance fitness:**

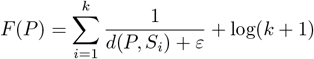

where *S*_*i*_ are siblings of node *P* and *d*(*P, S*_*i*_) represents the phylogenetic distance from node *P* to sibling *S*_*i*_.

- *Benefits:* Very sensitive to possible bottlenecks.
- *Drawbacks:* Does not consider possible descendants of node *P* .

- **Root-to-leaf distance fitness:**

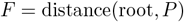

measuring evolutionary depth of node *P* .

- *Benefits:* Reflects lineage persistence or survival time.
- *Drawbacks:* Does not have global context.

- **Combined fitness:**

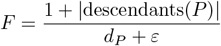

integrating both subtree size and terminal branch length.

- *Benefits:* Balances short-term evolutionary rate with long-term lineage success.
- *Drawbacks:* More complex; sensitive to parameter *ε* choice affecting numerical stability.

- **Child Distance Fitness:**

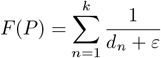

- *Benefits:* Captures the fine-grain impact of each child.
- *Drawbacks:* Nodes with fewer but more distant children of higher importance can be undervalued, also much more computationally intensive.

## 9 Fitness Visualizations

## 10 Fitness Metric Analysis

The following is based on the histograms shown in Figures 17–24, including bar graphs of fitness distributions and scatter plots comparing fitness to various traits.

### 10.1 Bar Graphs of Fitness Distributions

- **Terminal Fitness (Figure 13):** The graph peaks near one, consistent with most nodes having short terminal branches. Frequent recent branching or efficient mutation accumulation could be the cause of these peaks.
- **Subtree Size Fitness (Figure 14):** The distribution is sharply peaked near zero, with a rapid drop-off. A minority of nodes represent large subtrees, which could be because few mutations achieve extensive proliferation.
- **Imbalance Fitness (Figure 15):** The fitness values are right-skewed again. Most nodes are situated in balanced tree regions, with only a subset lying in highly asymmetric clades that would score highly in this method of finding fitness.
- **Root Distance Fitness (Figure 16):** A multimodal pattern emerges, most likely due to two separate time-domain outcroppings of the simulated viruses. In other words, peaks at different distances from the root suggest distinct waves of diversification.
- **Combined Fitness (Figure 17):** The distribution is sharply peaked near zero, with a rapid drop-off which resembles Figure 14.
- **Child Distance Fitness (Figure 18):** The distribution is sharply peaked near zero, with a rapid drop-off. This reflects that most nodes have few close descendants, while a small number are key to diversification events.

**Figure 13:**
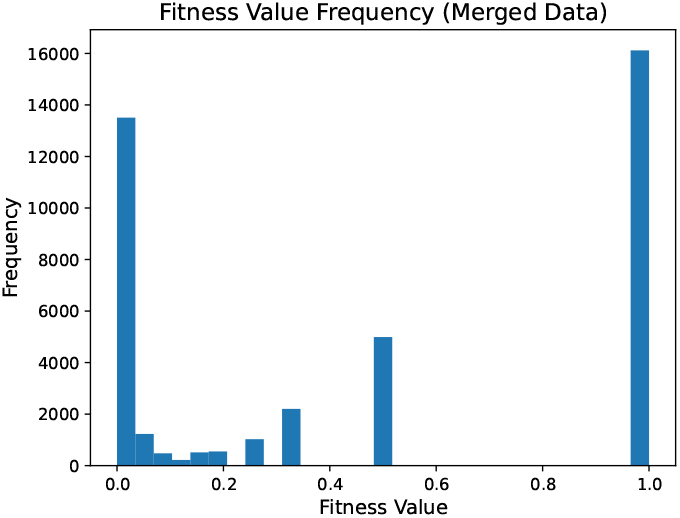
Terminal Fitness Bar Graph.

**Figure 14:**
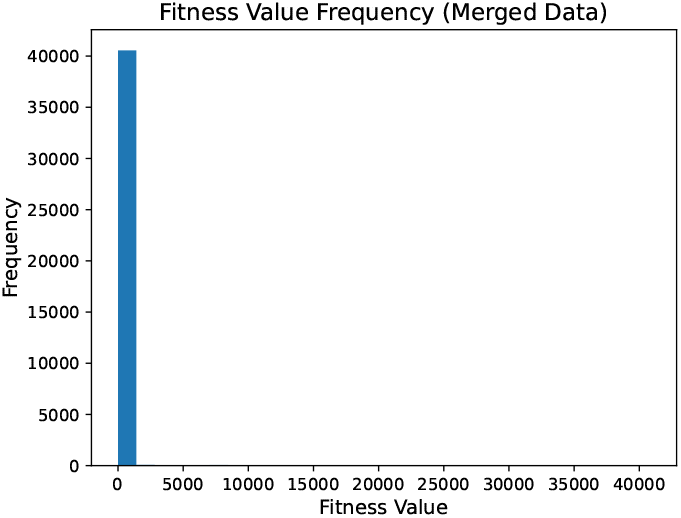
Subtree Fitness Bar Graph.

**Figure 15:**
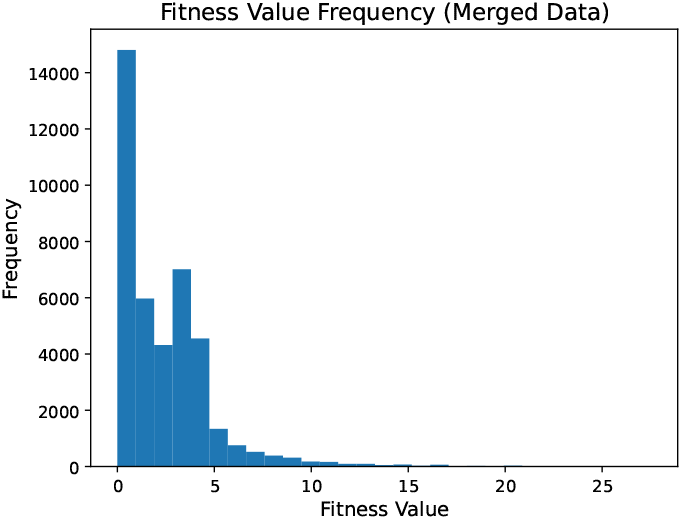
Imbalance Fitness Bar Graph.

**Figure 16:**
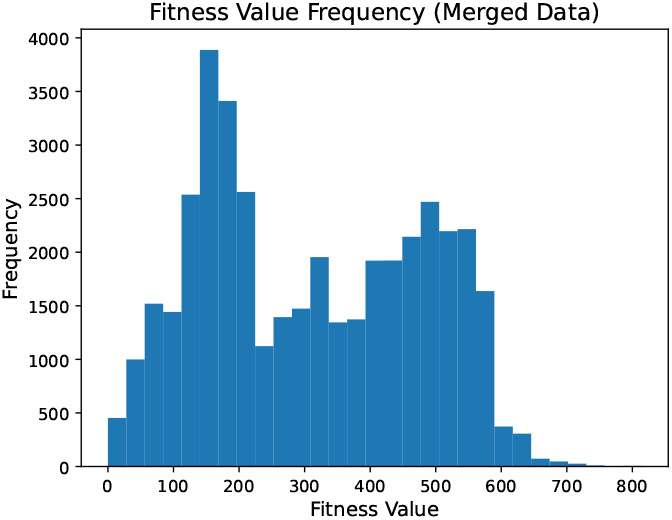
Root Distance Fitness Bar Graph.

**Figure 17:**
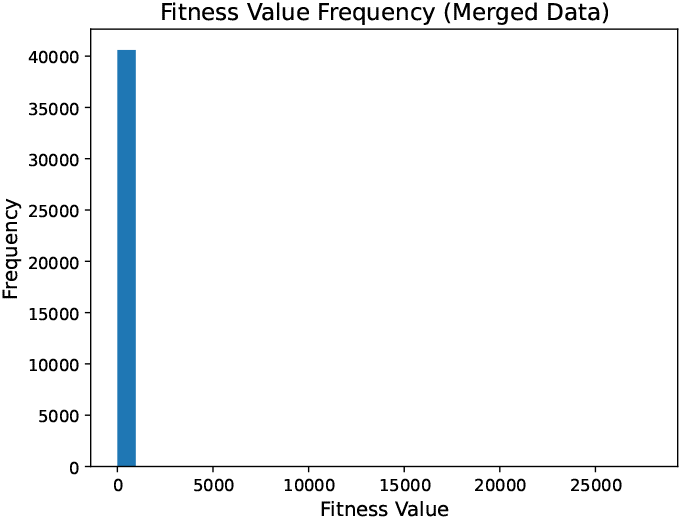
Combined Fitness Bar Graph.

**Figure 18:**
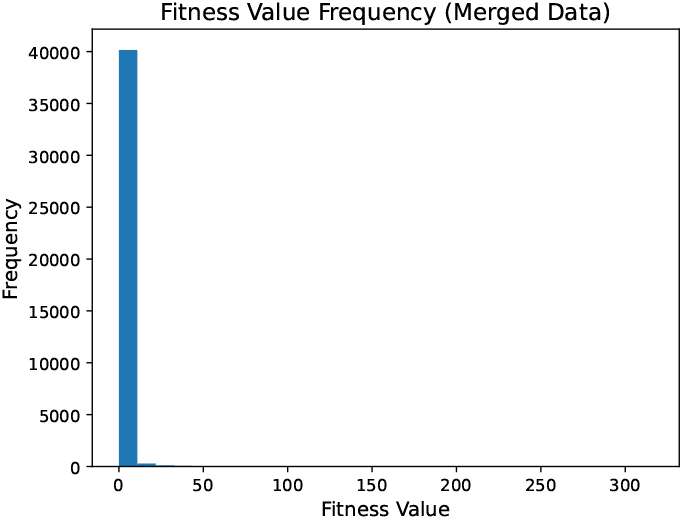
Child Distance Fitness Bar Graph.

### 10.2 Scatter Plots: Fitness vs. Traits

- **Terminal Fitness (Figure 19):** Displays high scatter and minimal trend lines, implying terminal branch length inverses do not capture trait change well.
- **Subtree Fitness (Figure 20):** Very limited correlation is observed across most traits, suggesting that total descendant count reflects expansion rather than any concrete trait.
- **Imbalance Distance Fitness (Figure 21):** Some linear trends are visible, indicating traits may partially track with diversification events. However, the relationships are extremely, extremely noisy.
- **Root Distance Fitness (Figure 22):** Some linear trends are visible, indicating traits may partially track with node age or survival time. However, the relationships are extremely, extremely noisy.
- **Combined Fitness (Figure 23):** Very limited correlation is observed across most traits, suggesting that the combined metric fails in the same way that the terminal metric or subtree metric fail.
- **Child Distance Fitness (Figure 24):** Very limited correlation is observed across most traits, suggesting that the child distance fitness metric may also fail to correlate to traits.

**Figure 19:**
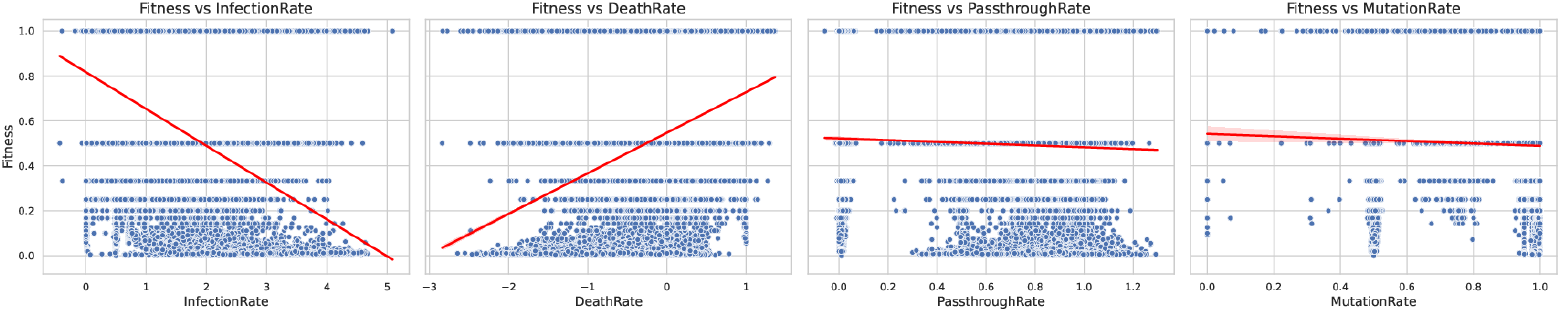
Terminal Fitness vs Traits.

**Figure 20:**
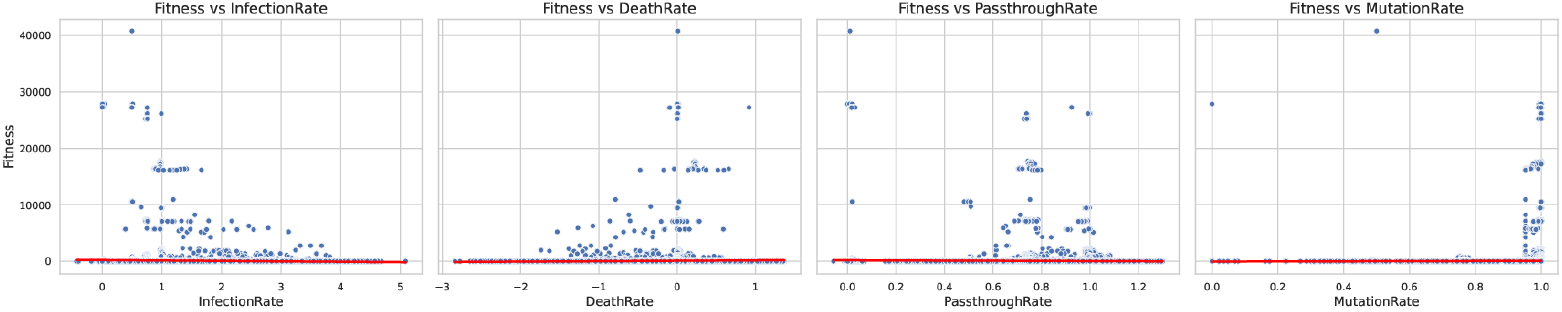
Subtree Fitness vs Traits.

**Figure 21:**
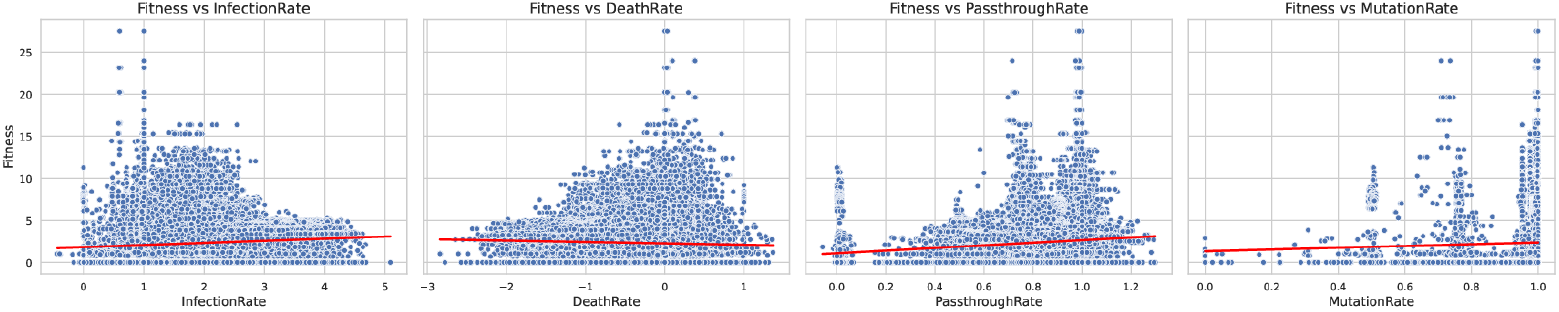
Imbalance Fitness vs Traits.

**Figure 22:**
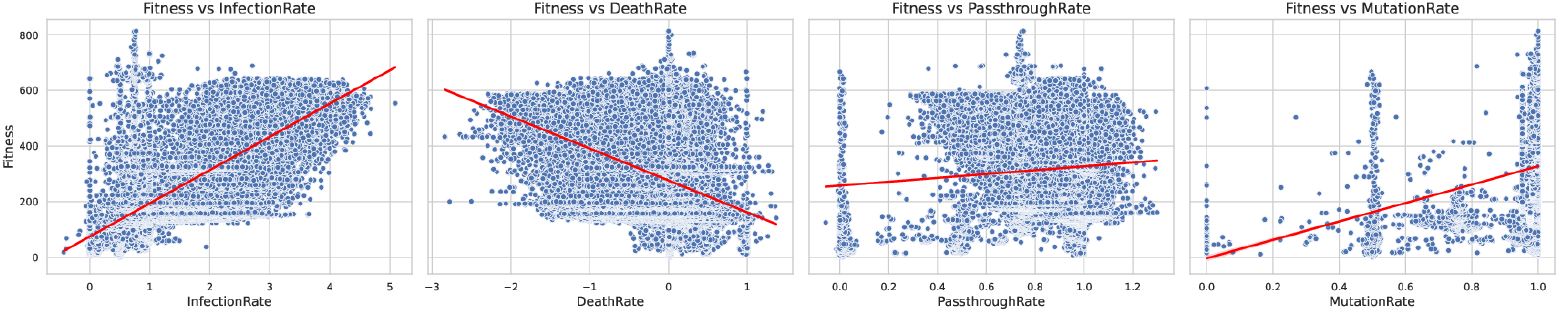
Root Distance Fitness vs Traits.

**Figure 23:**
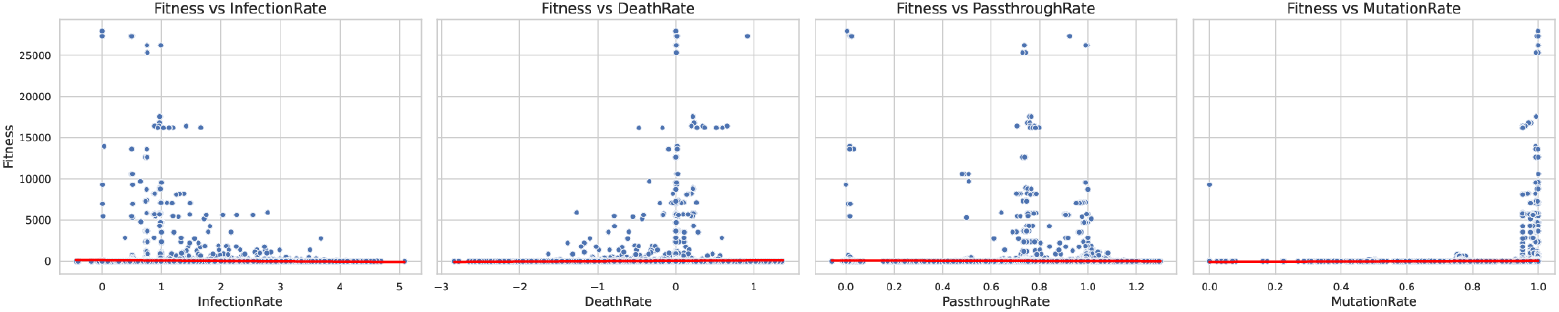
Combined Fitness vs Traits.

**Figure 24:**
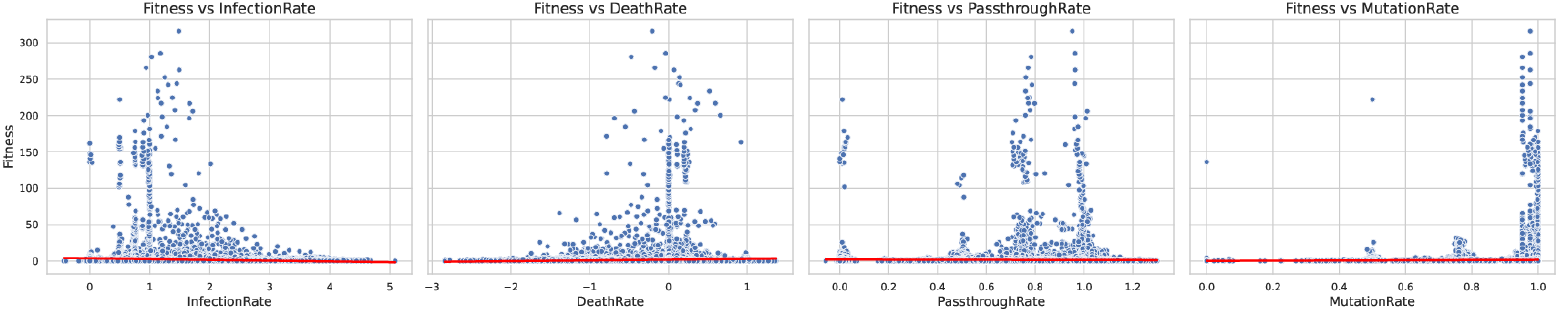
Child Distance Fitness vs Traits.

### 10.3 Summary

- **Distributional Skew:** All fitness metrics show skewed distributions, with most nodes concentrated near zero and fewer nodes attaining high scores. This aligns with biological expectations of uneven reproductive success and lineage survival.
- **Trait Association:** Combined and root distance metrics demonstrate some alignment with trait variation, implying utility in comparative analyses. Other metrics (terminal, subtree, child distance) primarily reflect structural properties of the tree without strong phenotypic correspondence. However, these trends are definitely not statistically significant, leaving much room for improvement in linking fitness with individual traits in the simulation.
- **Possible Causes:** Different fitness metrics illuminate distinct aspects of viral evolution. Structural metrics are useful for tree shape analysis, while combined metrics may serve as more effective proxies for actual biological fitness when phenotype data are available. More than that, individual traits on their own may not hold enough weight to fully correlate with fitness. Instead, more likely, is that the connection between differing traits that correlates with fitness.

## 11 Conclusion

`ViralSim` presents a flexible, geographically grounded simulation framework for modeling viral transmission, mutation, and evolution. By focusing on agent-based dynamics across spatially defined clusters, the system offers a realistic yet controllable platform for studying how viruses propagate and diversify. The incorporation of multiple phylogeny-based fitness metrics adds analytical depth, enabling researchers to quantify viral lineage success from structural features of the tree alone. Together, these components make `ViralSim` a useful tool for testing hypotheses about pandemic formation, evaluating reconstruction algorithms, and exploring the relationship between trait evolution and viral fitness.

## 12 Next Steps

Future development should prioritize enhancing computational efficiency, focusing on both execution speed and memory optimization to enable running much larger simulations. Expanding customization options and implementing a broader suite of commands will increase the flexibility and applicability of the simulation framework. For example, greater customization in the total number of traits could result in more complex equations connecting the traits together. While more complexity does not equate to a better model, having that option is nonetheless useful.

Additionally, rigorous validation of fitness metrics against empirical data is essential. The purpose of estimating fitness is to eventually apply fitness to realworld viruses – either by solely utilizing the phylogeny for fitness or by estimating trait values for real-world viruses. Also, a stronger fitness metric might result in more clear correlation through trait values. While not a huge concern at the moment, a predictive model may have better performance at predicting fitness from traits. Also, integrating established phylogenetic comparison metrics such as Robinson-Foulds distances or Branch Score differences can provide more clear metrics at how simulation phylogenies (or the true tree) compare with reconstructed phylogenies (such as through maximum likelihood.

There is one more clear next step which should be taken: connecting geography with fitness. ViralSim’s uniqueness is in simulating geography. Thus, it seems counter-intuitive to not include that metric within the fitness analyses. However, at the same time, it’s difficult to imagine a concrete, scientifically-grounded idea that allows insights into fitness through geography.

## Data Availability

All data produced are available online at https://codeberg.org/melthorm/ABMTC and nextstrain.org

## 13 Code and Data Availability

The full source code used for simulation, fitness computation, and downstream analysis is available at: https://codeberg.org/melthorm/ABMTC.

Simulated datasets used for testing and benchmarking are located in the repository under: /ABMTC/ViralSim/tests/.

Real-world SARS-CoV-2 phylogenetic data were obtained from NextStrain and ran with IQ-Tree: https://nextstrain.org/.

All outputs from analysis pipelines, including merged trait and fitness CSVs, plots, and diagnostic results, are stored in: /ABMTC/analysis/output data/.

## 14 Acknowledgements

I’d like, first and foremost, to thank Emma Goldberg for her help throughout this entire project as well as with research in general. I’d also like to thank the Institute for Computing in Research for providing the conditions necessary to start developing ViralSim. I’d also like to thank Mr. Shockey’s Independent Study Class for their help with the various coding problems and segfaults.

